# Continuously-Encoded Deep Recurrent Networks for Interpretable Knowledge Tracing in Speech-Language and Cognitive Therapy

**DOI:** 10.1101/2020.11.08.20206755

**Authors:** Ehsan Dadgar-Kiani, Veera Anantha

**Affiliations:** Department of Bioengineering, Stanford University, Stanford, CA 94305; Constant Therapy Health, Newton, MA 02458

## Abstract

Intelligent Tutoring Systems (ITS), developed over the last few decades, have been especially important in delivering online education. These systems use Knowledge Tracing (KT) to model a student’s understanding of concepts as they perform exercises. Recently, there have been several advancements using Recurrent Neural Networks (RNNs) to develop Deep Knowledge Tracing (DKT) that eliminates the need for manually encoding the student knowledge space. In online education, these models are crucial for predicting student performance and designing personalized curricula (sequence of courses and exercises). In this paper we develop a novel Knowledge Tracing model, called Continuously-encoded Deep Knowledge Tracing (CE-DKT) to automatically encode the user’s knowledge space, when the user’s skill in a given task is continuous-valued instead of binary. We then apply Knowledge tracing, specifically CE-DKT, to the context of digital therapy. Specifically, patients suffering from various neurological disorders such as aphasia, traumatic brain injury, or dementia are often prescribed speech, language and cognitive therapy exercises to perform from a set of predefined workbooks that are not personalized for the patient. We use CE-DKT to automatically encode a patient’s skill level across different tasks, and predict how the patient will perform on unseen tasks. We use data from the digital therapy platform, Constant Therapy, to train a CE-DKT model and demonstrate its high degree of accuracy in predicting a patient’s performance in a digital therapy application. We also demonstrate how to extract interpretable confidence intervals from this model and how to trace predictions to previous tasks using time-step level feature importance. Finally, we describe how this model can be applied to significantly enhance future digital therapy platforms and online student learning systems.

## 1 Introduction

Intelligent Tutoring Systems (ITS) have been developed over the last few decades and have been especially important in delivering online education [4]. In these systems, Knowledge tracing (KT) is the task of modeling a student’s understanding of concepts as they perform exercises, and this allows the prediction of student performance on future exercises. Even when the exact relationships between types of exercises are known, KT is intrinsically a challenging task because a student’s ability to learn a concept and transfer it to a different exercise is widely variable. Recently, there have been several advances using Recurrent Neural Networks (RNNs) to develop Deep Knowledge Tracing (DKT) models [11]. The DKT family of models automatically encodes the student’s ability/mastery with different tasks, thus eliminating the need for manually encoding the student’s knowledge space.

However, current DKT models developed thus far have a significant limitation in that they were only designed for binary inputs and outputs. Thus they cannot be used to model a continuous-valued representation of *how well* a patient is performing in their skills, such as is common in speech-language and cognitive therapy applications, and in many student learning applications.

In this paper, we make two broad contributions: (1) We improve upon the DKT model and develop the Continuously-Encoded Deep Knowledge Tracing (CE-DKT) model that is more interpretable and usable for a broader set of applications than was possible until now. (2) We then apply the CE-DKT model developed in this paper in a novel manner to the healthcare problem of digital speech, language and cognitive therapy.

## 2 Summary of Contributions and Model Improvements

The main improvements developed in our CE-DKT model compared to earlier DKT models are as follows:

1. We developed a novel data encoding approach for RNNs used in knowledge tracing, that can work well with continuous-valued inputs. Thus the CE-DKT model can be used in any online learning system that outputs continuous-valued scores for user task performance. This is an improvement over current DKT models that can only work with binary (correct/incorrect) inputs.
2. We demonstrate the application of CE-DKT to create a more computationally efficient knowledge tracing model, where the input at each time-step of the model can be the continuous-valued scores generated after the user has completed several exercises of a given type. This is an improvement over current DKT models that require an input after the user completes every exercise.
3. We developed an improved and more interpretable loss function, which directly represents the prediction error.
4. We demonstrate that our GRU-based model outperforms previous iterations of LSTM DKT variants.
5. We demonstrate the application of CE-DKT to the problem of digital speech-language and cognitive therapy, so that one can automatically track a patient’s knowledge / skill across different tasks and predict their future performance.

## 3 Related Work

There have been several decades of development of Intelligent Tutoring Systems and Knowledge Tracing models, the majority of which have been tested on online education (learning) data, and not on healthcare (digital therapy) data, which is the subject of this paper. Nonetheless, we will briefly mention several of the most notable approaches here.

The introduction of Deep Knowledge Tracing by Piech et al [11] demonstrated the first application of deep recurrent networks to student knowledge modeling, reporting substantial improvements in accuracy when compared to more established techniques such as Bayesian Knowledge Tracing (BKT) [11]. Subsequent studies since then by others in the field have incorporated additional high-dimensional features into the input vector [14], incorporated regularization for accurately predicting previous time steps and reducing the waviness of the prediction [12], incorporating both attention, transformer, and memory networks for improved interpretability of the model’s underlying relationships between concepts [13].

Most previous studies relied on either synthetic data or the publicly-available ASSISTment dataset^1^, which contains longitudinal student performance across various skill sets. This dataset is quite limited, in that it only contains a binary representation of how a student performed on a given exercise. As such, these models were developed for applications to feed-in and predict binary input/output. This may be sufficient for applications where it is adequate to know if a student either understands / does not understand (binary) a concept such as “multiplication” or “division”, but it is not useful when we want to know *how well* a student understands such concepts. For the general student learning application and digital therapy applications, such as where one must assess a patient’s brain recovery after a stroke or traumatic-brain-injury, it is important to know how well the patient understands these concepts, and thus a more continuous-valued representation of the student / patient’s knowledge is required. The CE-DKT model developed in this paper provides such a generalized approach that can work well in these applications.

## 4 Model

In this paper we develop the Continuously-Encoded Deep Knowledge Tracing (CE-DKT) model that uses a Recurrent Neural Network (RNN). RNNs are very useful in applications where a sequence of inputs is used to predict a sequence of outputs [5]. In this application, the sequence of inputs is defined by the users’ performance (accuracy scores) on the sequence of exercises they perform over time. At each time-step *t*, the model predicts the user’s expected performance (accuracy score) on all possible exercises they could do in the next time-step *t* + 1); this output prediction can be interpreted as the user’s expected “skill level” in any exercise they choose to perform next. Figure 1.a shows the original DKT model, while Figure 1.b shows the CE-DKT model developed in this paper. Next we describe this model in detail.

**Figure 1:**
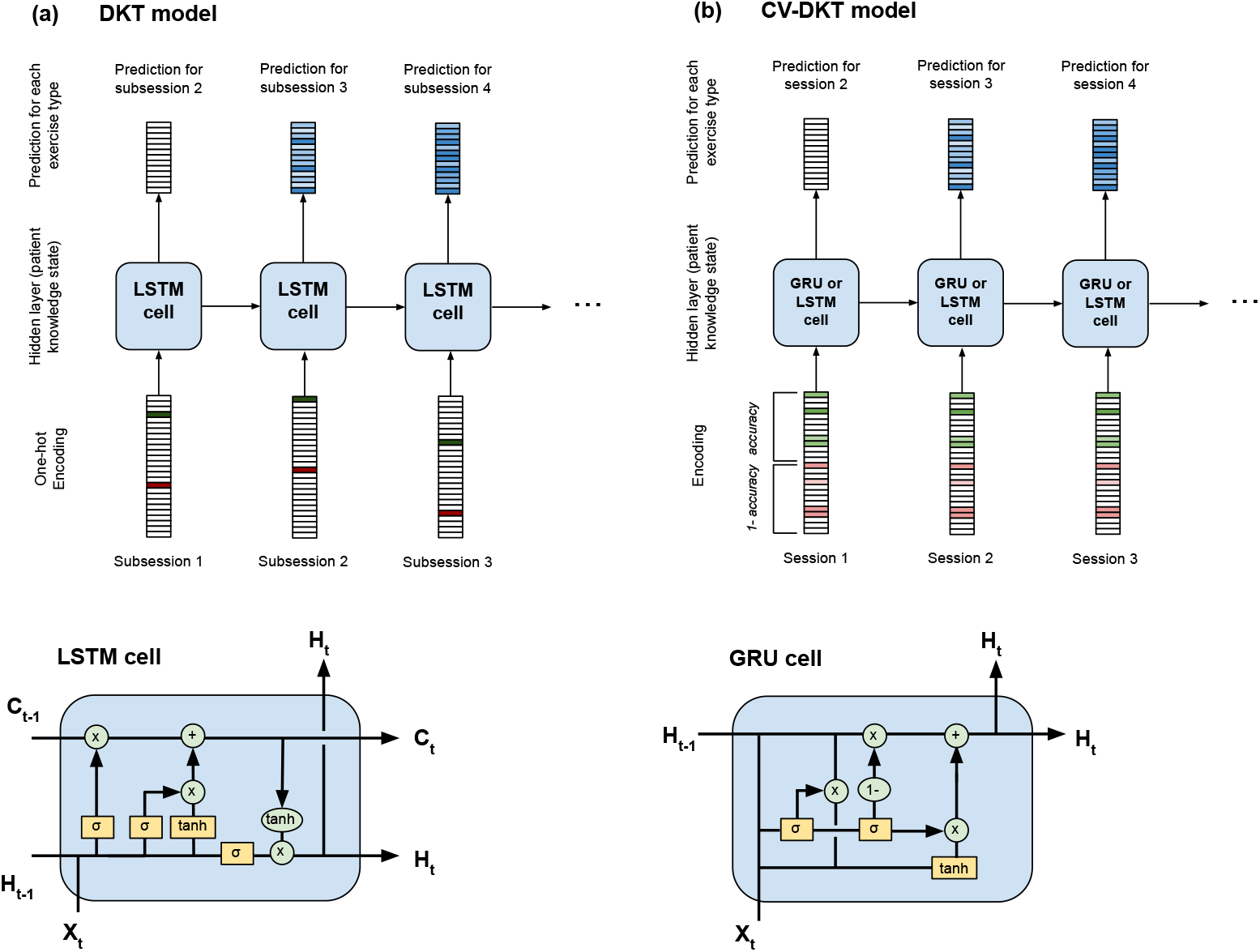
We depict both the original DKT model [11] that uses a one-hot binary encoding of accuracy, and the CE-DKT model developed in this paper that encodes multiple continuous accuracies. 1.a: Shows the original DKT model with LSTM recurrent network. 1.b: Shows the CE-DKT model variant that uses the GRU recurrent network. 1.c: Shows the structure of a standard LSTM cell. 1.d: Shows the structure of a standard GRU cell.

### 4.1 Data Encoding

The original DKT implementation by Piech [11] (shown in Figure 1.a) had proposed a *binary* one-hot encoding mechanism *x* ∈ {0, 1}^2*M*^, where *M* is the number of exercise *types*. The first half of the vector *x* represents whether the user answered the exercise correctly, while the second half represents whether they answered this exercise incorrectly.

The CE-DKT model is shown in Fig 1.b, and it enhances the encoding approach of DKT in 2 significant ways. First, we use a *continuous-valued* score instead of a *binary* score to represent how well the user performed on a given task. This generalization is crucial in digital therapy and student learning applications as it allows us to handle a much larger class of exercise types, where the user’s performance score is not a simple binary number (right / wrong). Thus the input vector in CE-DKT is *x* ∈ [0, 1]^2*M*^ instead of *x* ∈ {0, 1} ^2*M*^. The first half of this vector contains the raw accuracy scores for the exercise type the user completed (and 0 if they did not perform that type of exercise), while the second half of this vector contains 1*− accuracy* for that type of exercise (and 0 if they did not perform that type of exercise).

The second significant enhancement to the encoding developed in the CE-DKT model is to *time-compress* multiple set of exercises performed by the user in a “*session*” into a *single* input vector *x*_*t*_ corresponding to *session t*. This is especially useful in digital therapy applications (or student learning applications) where a patient (student) typically performs a prescribed sequence of therapy exercises in one “sitting” or “therapy session”. The encoding model we use in CE-DKT creates the *time-compressed* encoded input vector *x*_*t*_ using the following approach: (1) If the user performs multiple exercises of a given *type* contiguously (defined as a *sub-session*), we compress these exercises into a *single* input vector. We calculate the *mean* accuracy score for all these exercises of a given *type* performed contiguously by the user (*sub-session* accuracy) - to create a single input vector *x*_*t*_ for the *sub-session t*, instead of multiple input vectors, one for each exercise completed. (2) If the user completes multiple *independent*^2^ *types* of exercises (multiple sub-sessions) in a single “sitting” or “session”, then we further compress all “sub-sessions” into a single input vector. Specifically, the mean accuracy scores for all distinct exercise types the user performed during this “session” (*t*) is included in a single input vector *x*_*t*_.

Using the CE-DKT encoding method, the input vector *x*_*t*_ is less sparse (not one-hot) compared to previous DKT approaches. Conversely, the number of time steps in the input sequence is significantly lower, allowing CE-DKT to more easily model applications where there are long sequences with long-term relationships between different elements of the sequence. This is especially true in the digital therapy (or student learning) applications, where an entire patient’s history can consist of upwards of hundreds of thousands of exercises that we would like to use to make a prediction. Overall, the CE-DKT encoding approach significantly improves the computational efficiency of the model without sacrificing the time sequence fidelity.

### 4.2 Architectures

In general, RNNs have known issues with carrying information across many time steps (from earlier to later), as well as the vanishing gradient problem during back-propagation (gradient shrinks too much as it is back-propagated through time) [10]. This is especially a problem in the digital therapy application considered in this paper, where an entire patient’s history can consist of upwards of hundreds of sessions, and we expect the model to capture short and long-term relationships between different elements of the sequence. Thus, in the CE-DKT model developed in this paper we explore the following variants of RNNs: Long Short Term Memory networks (LSTM) and Gated Recurrent Units (GRU).

LSTM (Long-short-term-memory) networks are one of the most widely used RNN variants that are able to retain information within a “cell state” across many time steps (long sequences) [5]. This is crucial for knowledge-tracing applications, since it is important to track the influence of a user’s current performance on their future performance many time steps from now. In the digital therapy application considered in this paper, the “cell state” can be interpreted as our representation of the patient’s “knowledge state” at the current point in time. In our application, this output represents our prediction of how the patient will perform on all of the different exercise types, given their current knowledge state. The matrices (*W*_*f*_, *W*_*i*_, *W*_*C*_, *W*_*o*_) and intercept terms (*b*_*f*_, *b*_*i*_, *b*_*C*_) are all trainable parameters that are optimized during the learning process. One important hyperparameter that must be properly set for all LSTM models is the dimensionality of the cell state.

Although most previous DKT studies have utilized either RNN or LSTM architectures, we additionally explored a variation of the Long Short Term Memory, called the Gated Recurrent Unit (GRU) [2]. Despite lower complexity and not having an additional cell state that is passed between time steps, the GRU model has the advantage of yielding similar performance with higher computational efficiency.

### 4.3 Loss Function

In order to train the CE-DKT model, we defined a loss function that can penalize model parameters when predictions do not match the training dataset. We define *Q*_*t*_ ∈ {0, 1} ^*M*^ as a vector containing 1’s for all the exercises the patient performed in session *t*, or 0 otherwise. Similarly, we define *A*_*t*_ ∈ [0, 1]^*M*^ as another vector containing the task-accuracies the patient achieved for all the different exercises they performed in session *t*, or 0 for the tasks they did not perform in that session. Finally, the CE-DKT model’s output vector *H*_*t*_ at time step *t* is the model’s prediction of the patient’s task-accuracies in the *next* session *t* + 1 on all types of exercises. Below, we denote this prediction output as *Y*_*t*_ = *H*_*t*_. Our primary loss function at time step *t* compares the model’s prediction of the task-accuracies for the patient in session *t* + 1, with the actual patient performance (task-accuracies) in that session:

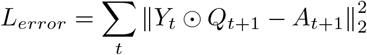

Here, ⊙ denotes element-wise multiplication. Thus for a given session at time *t, L*_*error*_ represents the model’s prediction accuracy (mean-squared-error) for session *t* + 1. We chose this as our primary loss-function because it makes the model more interpretable compared to previous models.

Previous models have proposed an additional regularization term to the loss function that attempts to simultaneously reconstruct the current and previous session’s (*t*) accuracies as well [12]. We also include these additional regularization terms in our loss function for CE-DKT.

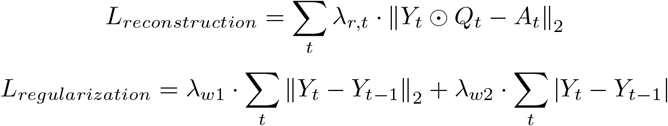

The hyperparameters *λ*_*r*_, *λ*_*w*1_, and *λ*_*w*2_ were selected using 5-fold cross-validation on the training set. Each model was trained using Stochastic Gradient Descent on the total loss function defined as:

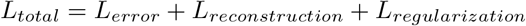

For this study, we used minibatch sizes of 100, with a learning rate of 0.01. Gradient explosion during backpropagation was prevented by clipping all gradients with norms greater than 5.0.

## 5 Dataset

The dataset for this study consisted of 8,713 patients who received speech, language and cognitive therapy through Constant Therapy, a mobile application for speech and language therapy developed by The Learning Corp. Each patient used the application for a minimum of 3 weeks, and although there were many patients who had used the application beyond 4 years, for the purposes of this study we disregarded a patient’s data beyond 2 years. At the time of the generation of this dataset, the Constant Therapy application supported 67 different exercise types across several cognitive and language categories. Each exercise type can potentially have many levels of difficulty, which brings the total count of exercise types (including different difficulty levels) to 309.

Patient performance on a given exercise is quantified as an accuracy score ranging from 0 to 1. For exercises that have multiple-choice questions, the score is binary (0 is incorrect, 1 is correct), whereas for more complex tasks, such as speaking or memory card tasks, the accuracy score represents the percentage of the exercise that the patient completed correctly. A patient typically performs multiple instances of an exercise type, for example 10 different multiplication exercises, which we collectively call a “sub-session”. For each “sub-session”, our dataset contains the mean accuracy and latency percentile scores (continuous valued). A patient typically performs a prescribed sequence of exercises for a day, which we collectively call a “session.” For each session, our dataset thus contains a list of mean accuracy and mean latency percentile values for all distinct exercise types they performed.

## 6 Results

We show the results for two primary variations of the CE-DKT model.

1. LSTM: In the case of the CE-DKT model that uses the LSTM cell at each time-step (session), we tried several variants of the LSTM cell with different cell state dimensions and found that increasing this hyper-parameter generally improves the model’s performance and decreases the test loss until saturation at about 1000. We did not exceed this value due to memory limitations during training.
2. GRU: In the case of the CE-DKT model that uses the GRU cell at each time-step. We found that this model, despite lower complexity, performs comparably as the LSTM models with largest cell state dimension.

First, we visualize the predicted performance (accuracy scores) of the CE-DKT model (GRU variant) for a given patient compared to the actual performance (accuracy score) for that patient. Figure 2 shows results for a patient from the Constant Therapy data set for Session 0 through Session 46. At each time step (i.e. therapy session), the CE-DKT model predicts the patient’s accuracy score on each type of exercise - this is represented by the background color of the square corresponding to that exercise type. As shown in the figure, for the first few sessions, the model’s predictions are not very good (it’s accuracy score prediction is uniformly close to zero). However after these initial sessions, the CE-DKT model’s predictions improve and can be used to predict the likely accuracy score the patient might get if they performed a given exercise type. The figure also illustrates that the model can be used to predict the patient’s performance on tasks they have never seen before, making the model very useful in practical applications.

**Figure 2:**
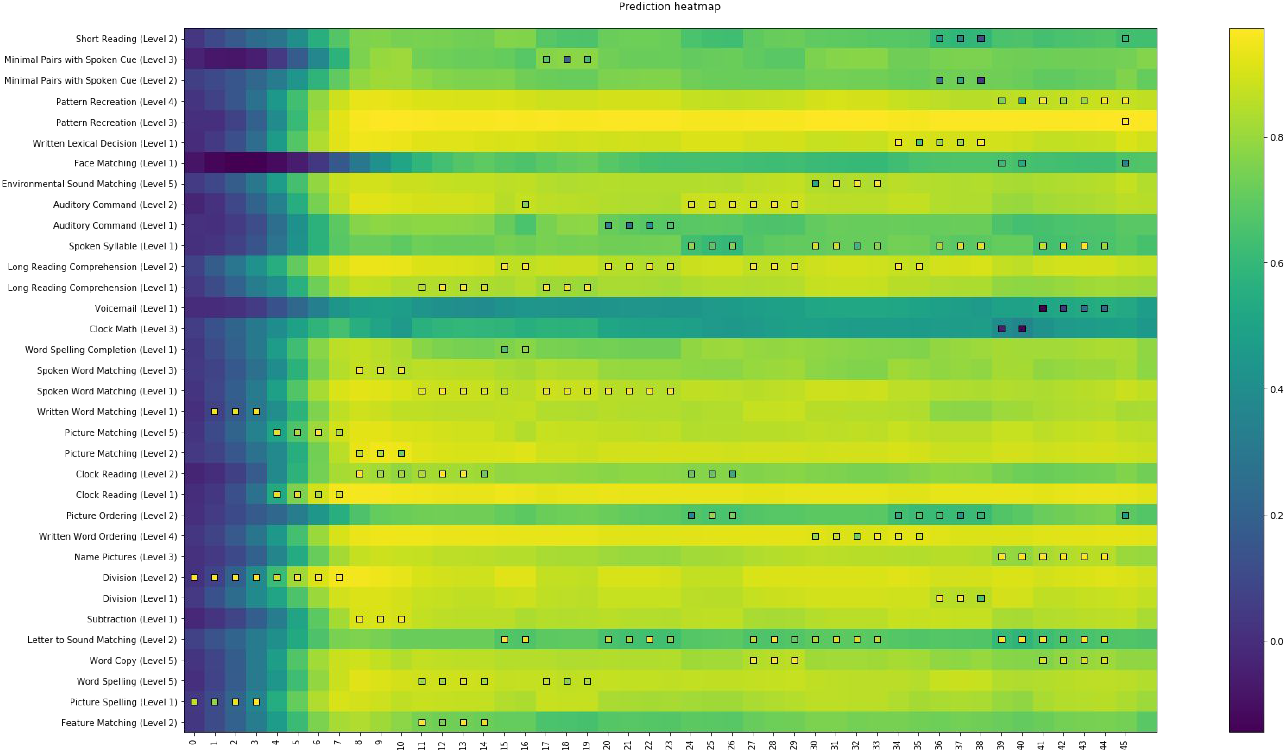
Visualizations of a patient’s full timeline of both actual and predicted performance. (a) Our heatmap representation of a patient includes the model’s prediction for all exercise types over time as the background color, while each foreground square represents a patient’s actual performance on an individual exercise at a certain time.

Next we quantify the performance of the different variants of the CV-DKT models. Assuming that the residuals between the predicted and actual variables have zero-mean, standard deviation *σ*, and are normally distributed, we can use the final loss value of the mean squared error to be the variance of the residuals. Additionally, taking the square root of this variance gives us an interpretable confidence interval with the same units as the output (accuracy percentage) to use alongside the model’s predictions. Thus, we also quantified the performance of our models using this more interpretable confidence metric (called “mean absolute prediction error”). Table 1 shows this for each variant of the model we implemented on the test data set. Based on these results, we conclude that the CE-DKT model that uses GRU cells performs either comparably or better than the various LSTM models, despite being a model with inherently lower complexity. Thus, we expect to use the GRU-variant of the CE-DKT model in future development and use.

**Table 1:** Model Results

| Model | Mean Abs. Error | Mean Abs. Error<br>(exercises with >1000 sessions) |
| --- | --- | --- |
| LSTM (10, cont.) | 0.3519 | 0.3435 |
| LSTM (50, cont.) | 0.3090 | 0.2949 |
| LSTM (100, cont.) | 0.3045 | 0.2879 |
| LSTM (500, cont.) | 0.2850 | 0.2721 |
| LSTM (750, cont.) | 0.2728 | 0.2572 |
| LSTM (1000, cont.) | **0.2714** | 0.2607 |
| GRU (w/ reg., cont.) | 0.2724 | **0.2510** |

We also explored the variation of the mean absolute prediction error for each exercise-type relative to the number of data-points (sessions) available in the training set for that exercise type. Figure 2-a shows the results of this relationship for the CE-DKT model variant that uses the GRU cells. As shown in the figure, we found a strong *negative* correlation (*r* = 0.56) between the mean-absolute-prediction-error for a given exercise type and the number of training data points for that exercise type. Thus the CE-DKT model’s predictions are much more accurate for the exercise types that have more training-data.

We additionally calculated the mean-absolute-prediction-error for each model variant, by excluding exercise types that had a relatively limited number of data points (less than 1000 total sessions) - results are summarized in the last column of Table 1. We see from these results that the mean-absolute-error is lower when we have more data to train the model (greater than 1000 total sessions); for example, the CE-DKT model that uses GRU cells has a mean-absolute-error of 0.25 while predicting the task-accuracy for tasks that have a training set of greater than 1000 sessions. We expect that as this model continues to be trained with even more data, the model prediction error will continue to go decrease even further.

This has practical significance, since the number of data points (sessions) will vary significantly across different exercise types in a typical training data set. For example, in the Constant Therapy data set we used in this paper, Aphasic stroke patients struggle to perform higher-difficulty-level speech-language exercises (so these exercise-types have fewer data points), while many more of these patients can do low-mid difficulty level speech-language exercises (thus they have more data points). When implementing the CE-DKT model in a practical “online” machine learning system, one could set a minimum threshold of the number of sessions of data needed in each exercise-type (for the Constant Therapy application it would be 1000 sessions), and only apply the model for those exercise types that have accumulated adequate training data. With such an approach, one can immediately start using the CE-DKT model, and over time all exercise types will have adequate training data to benefit from the model.

Finally, we also recalculated the model’s performance as we varied the number of first sessions we omitted in our error calculation, which we ranged from 0 to the first 60 sessions (Fig. 3-d). We found that the model takes approximately 10 sessions to be able to begin to accurately predict a patient’s performance. When only considering the exercise types with sufficiently large data set size (greater than 1000 total sessions), the mean error essentially saturates after about a patient’s first 10 sessions.

**Figure 3:**
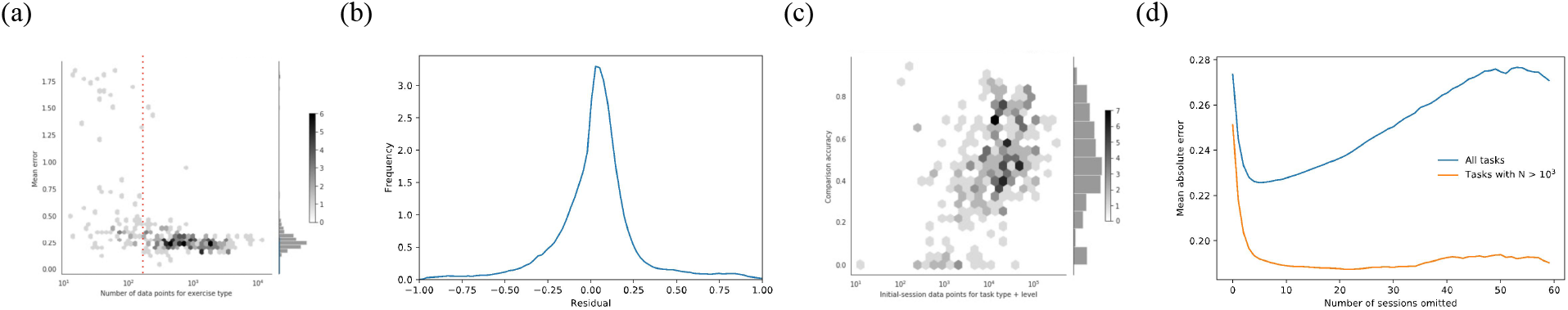
Performance for the best CE-DKT model. (a) For each exercise type, we plotted the model’s mean absolute error against the size of the training data set (in sessions). (b) Distribution of residual values between actual and predicted scores. (c) Our unseen-task metric plotted against training data set size, for each exercise type. (d) The model’s mean absolute error plotted against the number of initial sessions that are ommitted in the error calculation; for the exercise types that were deemed to have an “acceptable” number of data points in the training data set (N > 1000), the error stabilizes to a very low 0.175.

### 6.1 Model Interpretability

Although we optimized the mean squared error, the mean absolute error is a more interpretable distance measure of the mean residual difference between the predicted and actual accuracies. Under the assumption that these residuals are independently and identically distributed, and follow a normal distribution, this provides us with a confidence interval to use for each exercise-type. The bounds of the 68-percent confidence interval based on the standard deviation, for example, is given by:

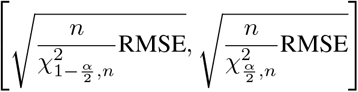

Such a confidence interval is crucial for a clinician or an automated system to evaluate the quality of this model’s prediction before using it to make a decision. For instance, if the model predicts extremely low performance with a high degree of accuracy, a clinician may choose to prescribe a lower level exercise, as to not demotivate the patient with an exercise that is too difficult.

Ultimately, a DKT model is most useful when predicting performance on exercise types the patient has yet to ever see. For this reason, we evaluate our model with an additional metric called the “unseen-task metric,” which we define as the percentage of times that our model is a better predictor of the first-session accuracy when compared to the population mean of all first-time sessions for that exercise-type. Figure 3(c) plots this metric across the different model types. We noticed a strong positive correlation (*r* = 0.71) between these two variables, again indicating that increasing orders of magnitude for each exercise type primarily improves the model’s performance for that exercise type.

## 7 Discussion

In this paper we demonstrated a novel application of Deep Knowledge Tracing specifically suited for the prediction of patient performance in speech-language and cognitive therapy applications. This required a new model, CE-DKT, that has the ability to simultaneously encode both the combination of exercises a patient performed in a given session as well as a continuous-valued representation of how they performed. We expect that this type of model would be useful in educational applications as well, especially as virtual learning is progressing and capturing higher dimensional interactions beyond just “correct” and “incorrect.” However, even with a DKT model that can accurately predict a patient’s performance on various exercise types, one of the crucial hurdles before full integration into the healthcare system is model interpretability and how one can use the model to influence dynamic decision making processes. Natural extensions to this model include incorporating the attention and transformer mechanisms into this sequence-to-sequence model, as these extensions have been previously shown to have significant improvements on knowledge tracing for educational applications [9].

### 8 Broader Impacts

A significant contribution of this paper is the application of Knowledge Tracing beyond adaptive student testing, learning and online curricula. Specifically we applied the CE-DKT model described in this paper to the healthcare problem of digital speech, language and cognitive therapy, which has many parallels to the problem of online student learning.

Patients suffering from neurological disorders such as traumatic-brain-injury (TBI), post-stroke aphasia, or dementia often have associated cognitive or language impairments such as difficulty with memory, speaking, problem-solving, or other cognitive skills. Clinical research has shown that these patients can regain their speech, language and cognitive skills by getting intense and long-term therapy [1][3][7][8]. Traditionally clinicians deliver such therapy via in-person sessions. However more recently, digital apps such as Constant Therapy [6] provide patients systematic therapy via their smartphones, tablets, or online. Patients getting “digital therapy” interact with exercises on their personal device, by responding to questions or problems presented to them. This is very similar to ITS where students respond to problems or questions presented to them online.

Although in traditional care, a clinician manually assesses a patient’s progress and decides what exercises to prescribe next, in digital therapy programs such as Constant Therapy, the system automatically identifies the right therapy exercise to present to each patient using a fixed set of rules based on the patient’s performance / progress^3^. These fixed rules are usually extrapolated from small scale clinical trials or population-based data analysis; these rules are manually encoded and provide a degree of personalization for each patient. The field of digital speech-language and cognitive therapy would benefit greatly from a system that can *automatically* encode the patient’s knowledge space, as that would lead to a much greater degree of personalized therapy for each patient. In this paper we explored the application of Knowledge Tracing to this problem, so that one can automatically track a patient’s knowledge / skill across different tasks and predict their future performance. Such a system would be useful for automated design of a custom curriculum (therapy regimen) for each patient to maximize their recovery.

Specifically, in this paper we applied the CE-DKT model to the digital therapy application and show the following:(1) We demonstrated how to use the CE-DKT model to trace the knowledge state of patients who are receiving speech, language and cognitive therapy through an online program. (2) We showed that this model can more accurately predict user performance on unseen tasks when compared to the population’s mean performance on these unseen tasks. (3) We demonstrated that the model’s predictive power improves with time as more exercises are processed by the model.

## Data Availability

Data is not publicly accessible via a public URL. However the data can be requested by contacting the Aphasia Research Lab at Boston University.

## Footnotes

1 https://sites.google.com/site/assistmentsdata/home/assistment-2009-2010-data

2 Exercise type A and B would be considered “independent” if the skills required to do exercise type A is independent of the skills required to do exercise type B.

3 https://www.constanttherapy.com

## References

[1] Mc Brady, H Kelly, J Godwin, P Enderby, and P Campbell. Speech and language therapy for aphasia following stroke (Review) SUMMARY OF FINDINGS FOR THE MAIN COMPARISON. Cochrane Database of Systematic Reviews, (6):2–7, 2016.

[2] Kyunghyun Cho, Bart Van Merriënboer, Caglar Gulcehre, Dzmitry Bahdanau, Fethi Bougares, Holger Schwenk, and Yoshua Bengio. Learning phrase representations using RNN encoder-decoder for statistical machine translation. EMNLP 2014 - 2014 Conference on Empirical Methods in Natural Language Processing, Proceedings of the Conference, pages 1724–1734, 2014.

[3] Keith D. Cicerone, Yelena Goldin, Keith Ganci, Amy Rosenbaum, Jennifer V. Wethe, Donna M. Langenbahn, James F. Malec, Thomas F. Bergquist, Kristine Kingsley, Drew Nagele, Lance Trexler, Michael Fraas, Yelena Bogdanova, and J. Preston Harley. Evidence-Based Cognitive Rehabilitation: Systematic Review of the Literature From 2009 Through 2014. Archives of Physical Medicine and Rehabilitation, 100(8):1515–1533, 2019.

[4] Arthur C. Graesser, Xiangen Hu, and Robert Sottilare. Intelligent tutoring systems. International Handbook of the Learning Sciences, 228:246–255, 2018.

[5] Sepp Hochreiter and Jürgen Schmidhuber. Long Short-Term Memory. Neural Computation, 1997.

[6] Swathi Kiran, Carrie Des Roches, Isabel Balachandran, and Elsa Ascenso. Development of an impairment-based individualized treatment workflow using an iPad-based software platform. Seminars in Speech and Language, 35(1):38–50, 2014.

[7] Natalie Kreitzer, Kelly Rath, Brad G. Kurowski, Tamilyn Bakas, Kim Hart, Christopher J. Lindsell, and Opeolu Adeoye. Rehabilitation Practices in Patients with Moderate and Severe Traumatic Brain Injury. Journal of Head Trauma Rehabilitation, 34(5):E66–E72, 2019.

[8] Aleksandra Kudlicka, Anthony Martyr, Alex Bahar-Fuchs, Bob Woods, and Linda Clare. Cognitive rehabilitation for people with mild to moderate dementia. Cochrane Database of Systematic Reviews, 2019(8), 2019.

[9] Shalini Pandey and George Karypis. A Self-Attentive model for Knowledge Tracing. 2019.

[10] Razvan Pascanu, Tomas Mikolov, and Yoshua Bengio. On the difficulty of training Recurrent Neural Networks. nov 2012.

[11] Chris Piech, Jonathan Spencer, Jonathan Huang, Surya Ganguli, Mehran Sahami, Leonidas Guibas, and Jascha Sohl-Dickstein. Deep Knowledge Tracing. 2015.

[12] Chun Kit Yeung and Dit Yan Yeung. Addressing two problems in deep knowledge tracing via prediction-consistent regularization. Proceedings of the 5th Annual ACM Conference on Learning at Scale, L at S 2018, 2018.

[13] Jiani Zhang, Xingjian Shi, Irwin King, and Dit Yan Yeung. Dynamic key-value memory networks for knowledge tracing. 26th International World Wide Web Conference, WWW 2017, pages 765–774, 2017.

[14] Liang Zhang, Xiaolu Xiong, Siyuan Zhao, Anthony Botelho, and Neil T. Heffernan. Incorporating rich features into deep knowledge tracing. L@S 2017 - Proceedings of the 4th (2017) ACM Conference on Learning at Scale, pages 169–172, 2017.

